# Childhood meningitis in rural Gambia: 10 years of population-based surveillance

**DOI:** 10.1101/2022.03.01.22271692

**Authors:** Usman N Ikumapayi, Philip C Hill, Ilias Hossain, Yekini Olatunji, Malick Ndiaye, Henry Badji, Ahmed Manjang, Rasheed Salaudeen, Lamin Ceesay, Richard A Adegbola, Brian M Greenwood, Grant A Mackenzie

**Author notes:** **Corresponding author:** Usman Nurudeen Ikumapayi, PhD, Senior Research Associate Disease Control and Elimination, Medical Research Council Unit at the London School of Hygiene and Tropical Medicine. **Alternate author:** Grant Mackenzie, Senior Clinical Epidemiologist and Principal Investigator, Disease Control and Elimination, Medical Research Council Unit at the London School of Hygiene and Tropical Medicine.

## Abstract

**Background:** The introduction in many countries of conjugate vaccines against *Haemophilus influenzae* type-b, *Streptococcus pneumoniae*, and *Neisseria meningitidis* has led to significant reductions in acute bacterial meningitis (ABM) in children. However, recent population-based data on ABM in sub-Saharan Africa are limited.

**Methods:** Population-based surveillance for meningitis was carried out in a rural area of The Gambia under demographic surveillance from 2008 to 2017, using standardised criteria for referral, diagnosis and investigation. We calculated incidence using population denominators.

**Results:** We diagnosed 1,599 patients with suspected meningitis and collected cerebrospinal fluid (n=1,121) and/or blood (n=1,070) from 1,427 (88%) of cases. We detected 169 cases of ABM, 209 cases of non-bacterial meningitis and 1,049 cases of clinically suspected meningitis. The estimated average annual incidence of ABM was high at 145 per 100,000 population in the <2-month age group, 56 per 100,000 in the 2–23-month age group, but lower at 5 per 100,000 in the 5–14-year age group. The most common causes of ABM were *Streptococcus pneumoniae* (n=44), *Neisseria meningitidis* (n=42), and Gram-negative coliform bacteria (n=26). Eighteen of 22 cases caused by pneumococcal serotypes included in PCV13 occurred prior to vaccine introduction and four afterwards. The overall case fatality ratio for ABM was 29% (49/169) and highest in the <2-month age group 37% (10/27). The fatality rate was 8.6% (18/209) for non-bacterial meningitis cases.

**Conclusions:** Gambian children continue to experience substantial morbidity and mortality associated with suspected meningitis, especially acute bacterial meningitis. Such severely ill children in sub-Saharan Africa require improved diagnostics and clinical care.

**Summary of the articles main point:** Population-based surveillance in a health demographic surveillance area in Gambia showed a high incidence and mortality in clinically suspected, acute-bacterial, and non-bacterial meningitis among children 14-years of age. Findings revealed potential gaps in the diagnosis of meningitis in The Gambia requiring urgent attention.

## Introduction

Acute bacterial meningitis (ABM) is an infectious disease syndrome characterised by bacterial infection and inflammation of the meninges, and is associated with high mortality and morbidity [1, 2]. ABM is among the major killers of children living in the meningitis-belt of sub-Sahara Africa [3], which still experiences high rates of endemic and epidemic meningitis due to *Neisseria meningitidis* [4, 5] *Haemophilus influenzae* type b (Hib) [6] and *Streptococcus pneumoniae* [7]. The recent introduction of conjugate vaccines in many African countries has led to some reductions in the incidence of meningitis due to Hib, *S. pneumoniae* and group A *N. meningitides* [7–9]. Given that most meningitis studies are hospital-based, there are limited population-based data on the incidence of meningitis in sub-Saharan Africa [10]. Furthermore, data on the incidence and outcome of non-bacterial meningitis in Africa are scarce.

In The Gambia, Hib and pneumococcal conjugate vaccines (PCV) were introduced in May 1997 (Hib), August 2009 (PCV7) and May 2011 (PCV13) respectively with high coverage (>90%). In 2013, individuals aged 1-29 years were vaccinated nationwide against the group A meningococcus [11]. Population-based surveillance has shown a substantial impact of the introduction of PCVs with an 80% reduction in the incidence of invasive pneumococcal diseases (IPD), in the under-5-year age group [12]. In this study, we used 10 years of standardised population-based surveillance for meningitis to describe incidence of meningitis over time, its aetiology, and case-fatality among children with ABM in a rural part of The Gambia in the era of widespread deployment of meningitis related vaccines (Hib, PCV and Men-A).

## Materials and Methods

### Study site and Participants

The Pneumococcal Surveillance Project (PSP) has conducted population-based surveillance for invasive bacterial disease in the Upper River Region of The Gambia since 2008 [13]. The PSP was based at the Basse Field Station of the Medical Research Council Unit The Gambia at the London School of Hygiene & Tropical Medicine (LSHTM).

We conducted surveillance for suspected pneumonia, sepsis and meningitis between May 12, 2008, and December 31, 2017. The population included all residents of the Basse Health and Demographic Surveillance System (BHDSS). The population was enumerated every 4 months, with births, deaths, migrations, and vaccinations been recorded. The estimated population in 2017 was 183,946 of whom 35,597 (20%) were younger than 5 years of age.

### Surveillance Procedures

Nurses used standardized criteria to screen all patients aged 2-59 months who presented at a health facility in the surveillance area for suspected septicaemia, pneumonia or meningitis, while infants in the first 2 months of life were screened only for suspected meningitis. Children who were positive on screening were referred to a clinician who used standardized criteria to determine a surveillance diagnosis and who initiated a standardized programme of investigation [14]. Meningitis in patients aged from 0-59 months of age was defined according to clinical judgement and suspected if one of the following was present: neck-stiffness, impaired consciousness, prostration, history of convulsion, or a bulging fontanelle. Among those aged 5 years and above, suspected meningitis was defined according to clinical judgement and considered if any of the following were present: axillary temperature 38°C and meningism (neck stiffness) and/or photophobia or altered mental state (Glasgow Coma Score <14). Patients with suspected meningitis had blood and cerebrospinal fluid (CSF) samples taken for culture. Inclusion criteria for this analysis were age ≤14 years and meeting the definition for suspected meningitis. Exclusion criteria were non-residence in the study area, trauma or a suspected hospital-acquired infection.

The definition for clinically suspected meningitis (CSM) cases was the presence of clinical features of meningitis, as described above, but a normal CSF leucocyte count 0-5/mm^3^ and a negative culture and antigen test. The definition of acute bacterial meningitis (ABM) was a CSF leucocyte count >5/mm^3^, and a positive culture of CSF or blood in a patient with clinical signs of meningitis. Non-bacterial meningitis (NBM), commonly termed aseptic meningitis, was defined as a CSF leukocyte count >5/mm^3^, a CSF negative antigen and a negative culture of CSF and blood in a patient with clinical signs of meningitis. All bacterial antigen positive CSF were also culture positive although some CSF had very scanty growth of <20 × 10^5^ organisms/mL. All bacterial antigen negative CSF were culture negative. However, only 87% of cultured CSF samples were tested for bacterial antigens.

### Outcomes

The primary outcome of the study was the incidence of acute bacterial and non-bacterial meningitis during the study period. Secondary outcomes were the aetiology of ABM and the case fatality rate in CSM, ABM, and NBM groups.

### Laboratory methods

Blood or CSF, or both, were collected for conventional microbiological investigations. Blood was inoculated into BACTEC bottles (Becton Dickinson) and incubated in an automated BACTEC 9050 blood culture system (Becton Dickinson) for a maximum of 5 days. Positive cultures were sub-cultured on blood and chocolate agar and examined for bacterial growth following 24 or 48-hours of aerobic incubation, incubation in 5-10% CO_2_ and anaerobic incubation, all at 37°C. Similarly, CSF was cultured on blood and chocolate agar and isolates identified by biotyping, and serotyping for pneumococcal serotypes. Cell count, Gram-stain and bacterial antigen (latex agglutination) tests were performed on CSF according to WHO procedures (WHO/CDS/CSR/EDC/99.7). Rapid diagnostic tests for malaria using ICT-Malaria *p.f*. Antigen tests (ICT Diagnostics) were performed routinely during the malaria transmission season. Children with clinical signs of meningitis but who had a positive rapid ICT diagnostic test for malaria and normal leukocyte count were excluded from our analysis.

### Antibiotic Susceptibility Testing

Disk diffusion methods were used to detect antibiotic resistance following standard guidelines (CLSI 2012, M100-S22, Vol. 32 No.3).

### Statistical analysis

Analyses were restricted to residents of the BHDSS. We calculated overall and age-stratified incidence for each category of patients during the observation period using overall and age-stratified numbers of cases divided by the overall and age-stratified BHDSS population denominators. We investigated trends in incidence over time, calculating annual incidence with mid-point population denominators from the BHDSS. We calculated disease incidence for three different categories of patient: CSM, ABM and NBM. Calculations of incidence and 95% confidence intervals assumed a Poisson distribution. We calculated case-fatality ratios (CFR) for each of the three categories of patient CSM, ABM and NBM based on the number of cases who died during and after hospital admission divided by the total number of cases in each category.

The period prior to the introduction of PCV13 (pre-PCV13) is defined as May 12, 2008 to December 31, 2012, whilst the period after the introduction of PCV13 (post-PCV13) is defined as January 1, 2013 until December 31, 2017. A p-value <0.05 was considered statistically significant.

### Ethical considerations

The study was approved by the Gambia Government-MRC Joint Institutional Ethics Committee (Re SCC 1087) and the ethics committee of the LSHTM. Guardian of participants gave written, informed consent for participation in the PSP.

## Results

### Demographic and clinical characteristics

Over the almost 10-year period of surveillance, 26,408 children were enrolled in PSP and 1,599 (6%) were referred by nurses for clinical review as possible cases of meningitis. Blood or CSF, or both, were taken from 1,427 (89.2%) of the suspected cases. The total number of blood cultures and CSF cultures performed were 1,070 and 1,121 respectively (Figure 1). Blood and CSF were obtained from 764 children, blood only from 306, and CSF only from 357. Approximately, 12% (169/1427) of suspected meningitis cases were bacterial antigen and culture positive, whilst bacterial antigen and culture negative cases accounted for 85% (1217/1427) and contaminants for a further 3% (41/1427) (Fig 1). Both ABM and NBM cases were more likely to be male than female. (Table 1).

**Figure 1:**
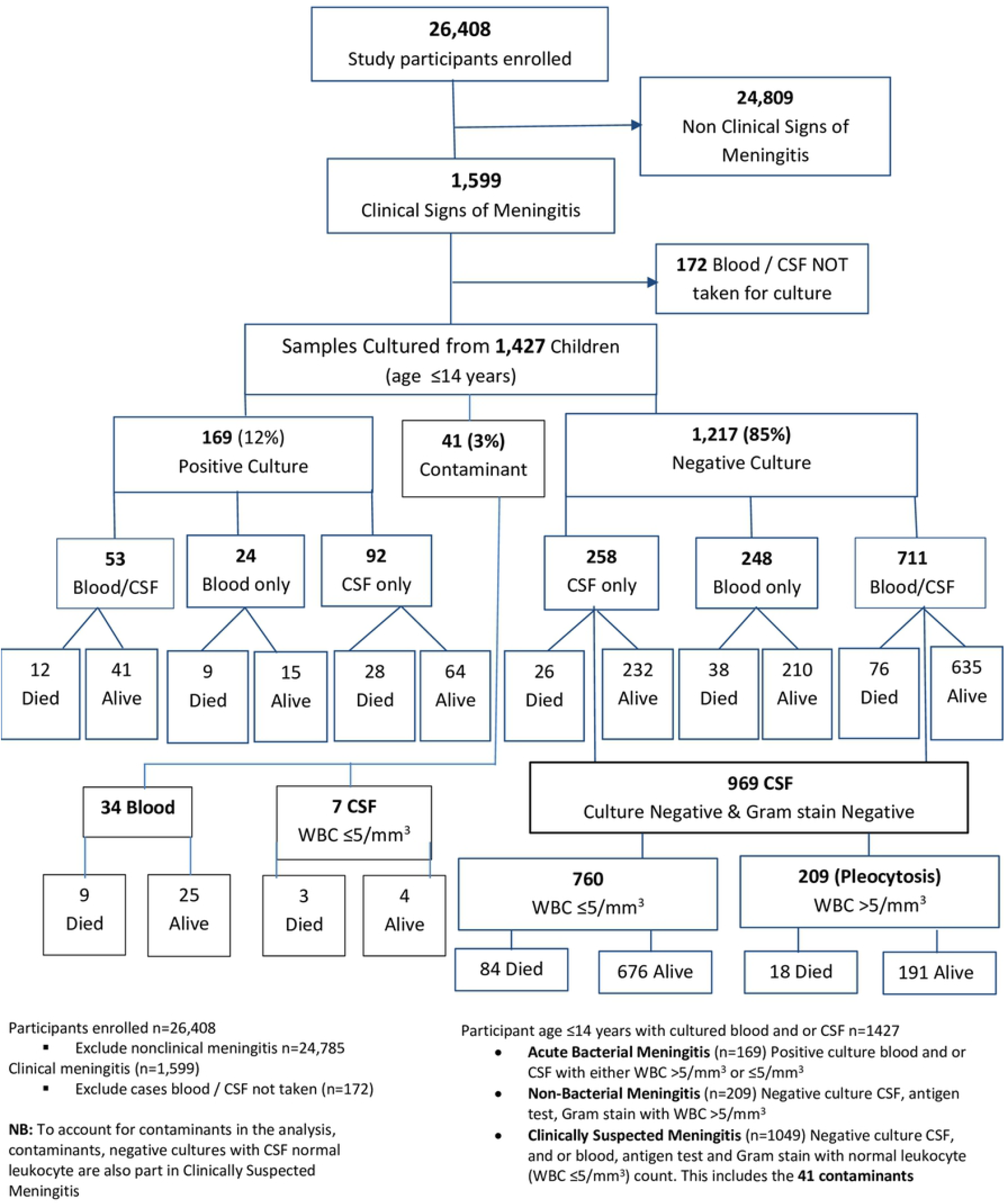
Enrolment, Diagnoses and Outcome:

**Table 1:** Baseline meningitis in rural Gambia: 10 years of population-based surveillance

| Characteristic | Category | Acute Bacterial<br>Meningitis (n=169)<br>N (%) | Non-Bacterial &<br>Clinically Suspected<br>Meningitis (n=1258)<br>N (%) | Total (n=1,427)<br>N | OR (95% CI) |
| --- | --- | --- | --- | --- | --- |
| Gender | Male | 90 (53.2%) | 734 (58.3%) | 824 | 0.81 (0.58-1.39) |
|  | Female | 79 (46.8%) | 524 (41.7%) | 603 | Reference |
| Age in months | <2 | 27 (50%) | 144 (55%) | 171 | 0.81 (0.43-1.53) |
|  | 2-23 | 65 (70.7%) | 488 (80.7%) | 553 | 0.58 (0.35-0.98) |
|  | 24-59 | 50 (64.9%) | 509 (81.3%) | 559 | 0.43 (0.25-0.74) |
|  | 60-168 | 27 (16.0%) | 117 (9.3%) | 144 | Reference |

| <u>Outcome</u> |  |  |  |  |
| --- | --- | --- | --- | --- |
| Outcome | Died (n=201)<br>N (%) | Survived (n=1,226)<br>N (%) | Total (n=1,427)<br>N | OR (95% CI) |
| ABM | 49 (24.4) | 120 (9.8) | 169 | 2.97 (1.99-4.37) |
| CSM | 134 (66.7) | 915 (74.6) | 1,049 | 0.68 (0.49-0.95) |
| NBM | 18 (9.0) | 191 (15.6) | 209 | Reference |

**Key:** ABM – Acute Bacterial Meningitis, NBM – Non-Bacterial Meningitis, CSM – Clinically Suspected Meningitis and N – Number

### Incidence

The incidence of ABM during the study period in all age groups combined was 20.3 per 100,000 (95% CI 17-24) (Table 2a and Table 2b) with a peak of 63.3 per 100,000 (95% CI 48-83) in 2012 (Table 2a). The incidence of ABM and NBM varied from year to year, while that of CSM was relatively stable over time. The incidence of clinically suspected meningitis and NBM was greatest in 2011, potentially related to an increase in presentation of convulsions associated with cerebral malaria and a high level of malaria transmission following widespread flooding in 2010 [14].

**Table 2a:** Incidence per 100,000 Population of Clinically Suspected Meningitis, Non-Bacterial Meningitis and Acute Bacterial Meningitis Among Children ≤ 14 Years of Age (2008-2017), By Year (n=1427)

| Year | Clinically Suspected Meningitis (n=1,049) |  |  | Non-Bacterial Meningitis (n=209) |  |  | Acute Bacterial Meningitis (n=169) |  |  |
| --- | --- | --- | --- | --- | --- | --- | --- | --- | --- |
|  | Cases | Incidence | 95% CI | Cases | Incidence | 95% CI | Cases | Incidence | 95% CI |
| **2008** | 97 | 279.5 | 227-341 | 12 | 34.6 | 18-60 | 8 | 23.1 | 10 - 45 |
| **2009** | 126 | 167.0 | 139-199 | 1 | 1.3 | 03-07 | 12 | 15.9 | 8 - 28 |
| **2010** | 122 | 154.7 | 128-185 | 13 | 16.5 | 09-28 | 16 | 20.3 | 12 - 33 |
| **2011** | 155 | 189.6 | 161-222 | 29 | 35.5 | 24-51 | 18 | 22.0 | 13 - 35 |
| **2012** | 101 | 118.4 | 96-143 | 35 | 41.0 | 29-57 | 54 | 63.3* | 48 - 83 |
| **2013** | 89 | 102.4 | 82-126 | 30 | 34.5 | 23-49 | 13 | 15.0 | 8 - 26 |
| **2014** | 112 | 127.1 | 105-153 | 22 | 24.9 | 16-38 | 23 | 26.1 | 17 - 39 |
| **2015** | 103 | 117.5 | 96-143 | 39 | 44.5 | 32-61 | 17 | 19.4 | 11 - 31 |
| **2016** | 77 | 87.4 | 69-109 | 17 | 19.3 | 11-31 | 4 | 4.5 | 1.2 - 12 |
| **2017** | 67 | 75.5 | 58-96 | 11 | 12.4 | 06-22 | 4 | 4.5 | 1.2 - 12 |
| **Total** | 1049 | 125.9 | 118-134 | 209 | 25.1 | 22-29 | 169 | 20.3 | 17 - 24 |

Note: Only 234 days of surveillance in 2008, from 12 May – 31 Dec.
\*Higher incidence due to the epidemic of *Neisseria meningitidis* W135

**Table 2b:** Incidence per 100,000 Population of Clinically Suspected Meningitis, Non-Bacterial Meningitis and Acute Bacterial Meningitis Among Children ≤ 14 Years of Age (2008-2017), By Age (n=1427)

| Age in Month | Clinically Suspected Meningitis (n=1,049) |  |  | Non-Bacterial Meningitis (n=209) |  |  | Acute Bacterial Meningitis (n=169) |  |  |
| --- | --- | --- | --- | --- | --- | --- | --- | --- | --- |
|  | Cases | Incidence | 95% CI | Cases | Incidence | 95% CI | Cases | Incidence | 95% CI |
| <2 | 115 | 616.6 | 509-740 | 29 | 155.5 | 104-223 | 27 | 144.8 | 95 -210 |
| 2-23 | 405 | 345.7 | 313-381 | 83 | 70.8 | 56-87 | 65 | 55.5 | 43 - 71 |
| 24-59 | 439 | 227.5 | 207-249 | 70 | 36.3 | 28-46 | 50 | 25.9 | 19 - 34 |
| 60-168 | 90 | 17.8 | 14-22 | 27 | 5.4 | 04-08 | 27 | 5.4 | 4 - 8 |
| **Total age** | 1049 | 125.9 | 118-134 | 209 | 25.1 | 22-29 | 169 | 20.3 | 17 - 24 |

## Case fatality

Mortality due to ABM was higher compared to NBM (OR: 2.97, 95% CI 1.99-4.37; (Table 1). Annual case fatality for ABM varied significantly from year to year whilst case fatality for CSM and NBM was relatively constant over time (Fig 2a). Overall case fatality was 29% (49/169) for ABM, 8.6% (18/209) for NBM and 12.8% (134/1049) for CSM. Mortality was highest in the first 2 months of life (37%), but also high in other age groups; 29%, 30%, and 18% in the 2-23 months, 2-4 years, and 5-14 years age strata respectively (Fig 2b). In all age strata, case fatality was lower for clinically suspected but not proven meningitis and for NBM compared to ABM (Fig 2b).

**Figure 2a:**
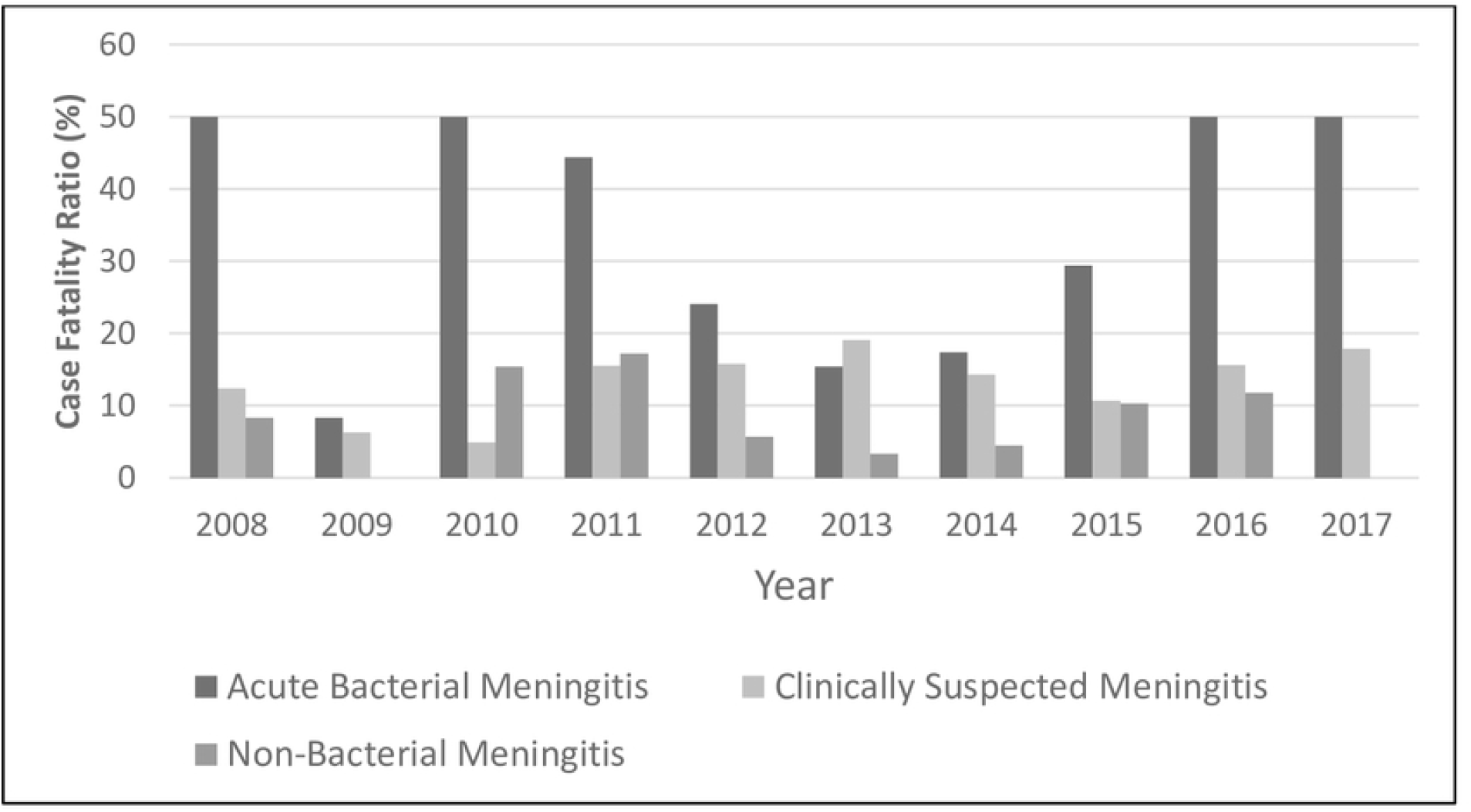
Annual Case Fatality Ratio of Clinically Suspected Meningitis (CSM), Non Bacterial Meningitis (NBM) and Acute Bacterial Meningitis (ABM) among children aged 1 day-14 years in Upper River Region Gambia, 2008-2017

**Figure 2b:**
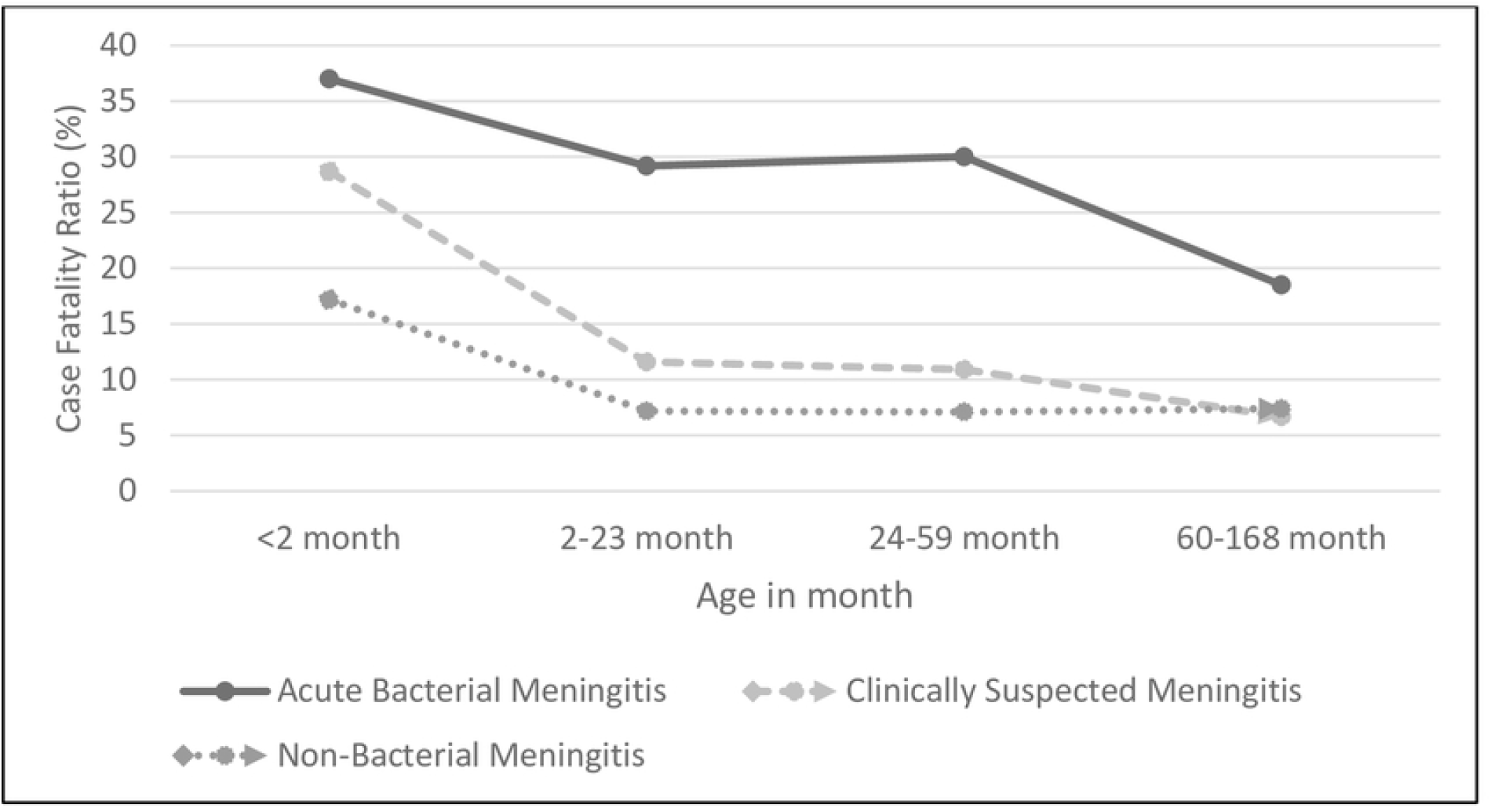
Age strata Case Fatality Ratio of Clinically Suspected Meningitis (CSM), Non Bacterial Meningitis (NBM) and Acute Bacterial Meningitis (ABM) among children aged 1 day-14 years in Upper River Region Gambia, 2008-2017

### Aetiology

Pathogenic bacteria were isolated from 77 blood cultures and from 92 CSF specimens from patients with a raised WBC count (Table 3). *S. pneumoniae* was isolated in 44 cases. *H. influenzae* type-b was found in 12 cases, 4 of whom had been vaccinated against Hib. There were 42 cases of meningococcal meningitis, all belonging to serogroup W, with 35 in 2012 during a W135 epidemic. Other common causes were Gram negative coliforms (n=26), *Staphylococcus aureus* (n=16) and non-typhoidal Salmonella (n=12), non-type b *H. influenzae* (n=6), *Klebsiella pneumoniae* (n=6) and *Escherichia coli* (n=5).

**Table 3:** Frequency of Bacteria isolated from blood and CSF cultured and corresponding case fatality ratio (CFR) caused among children in Upper River Region Gambia, 2008-2017 (n = 169)

| Bacteria | Total<br>N | Blood<br>N | CSF<br>N | No. of<br>Death | CFR<br>% | 95% CI |
| --- | --- | --- | --- | --- | --- | --- |
| **All Bacteria** | 169 | 77 | 92 | 49 | 29 | 21.4% - 38.3% |
| *Streptococcus pneumoniae* | 44 | 16 | 28 | 15 | 34.1 | 19.1% - 56.2% |
| *Neisseria meningitidis* W | 42 | 10 | 32 | 9 | 21.4 | 9.8% - 40.7% |
| Coliform (GNR) | 26 | 20 | 6 | 5 | 19.2 | 6.2% - 44.9% |
| *Staphylococcus aureus* | 16 | 13 | 3 | 6 | 37.5 | 13.8% - 81.6% |
| *H. influenzae* type b | 12 | 1 | 11 | 3 | 25 | 5.2% - 73.1% |
| Non-Typhoidal Salmonella | 12 | 8 | 4 | 4 | 33.3 | 9.1% - 85.3% |
| Non-Hib | 6 | 1 | 5 | 1 | 16.7 | 0.4% - 92.9% |
| *Klebsiella pneumoniae* | 6 | 6 | 0 | 4 | 66.7 | 18.2% - 170.7% |
| *Escherichia coli* | 5 | 2 | 3 | 2 | 40 | 4.8% - 144.5% |

*S. pneumoniae* was categorised into vaccine (n=22) and non-vaccine (n=22) serotypes based on the serotypes included in PCV13 (Table 4). We detected six vaccine serotypes (1, 5, 14, 6A, 19F and 23F). Eighty-two percent (18/22) of cases with vaccine serotypes were detected in the pre-PCV13 period compared to 18% (4/22) after the introduction of PCV13, one of whom had been vaccinated. We detected 15 non-vaccine serotypes (2, 21, 46, 9A, 10F, 12B, 12F, 15A, 15B, 16F, 17F, 18A, 23B, 25F and 35B), a half (11/22) of the cases infected with one of these non-vaccine serotypes were detected pre and post PCV 13 introduction respectively.

**Table 4:**
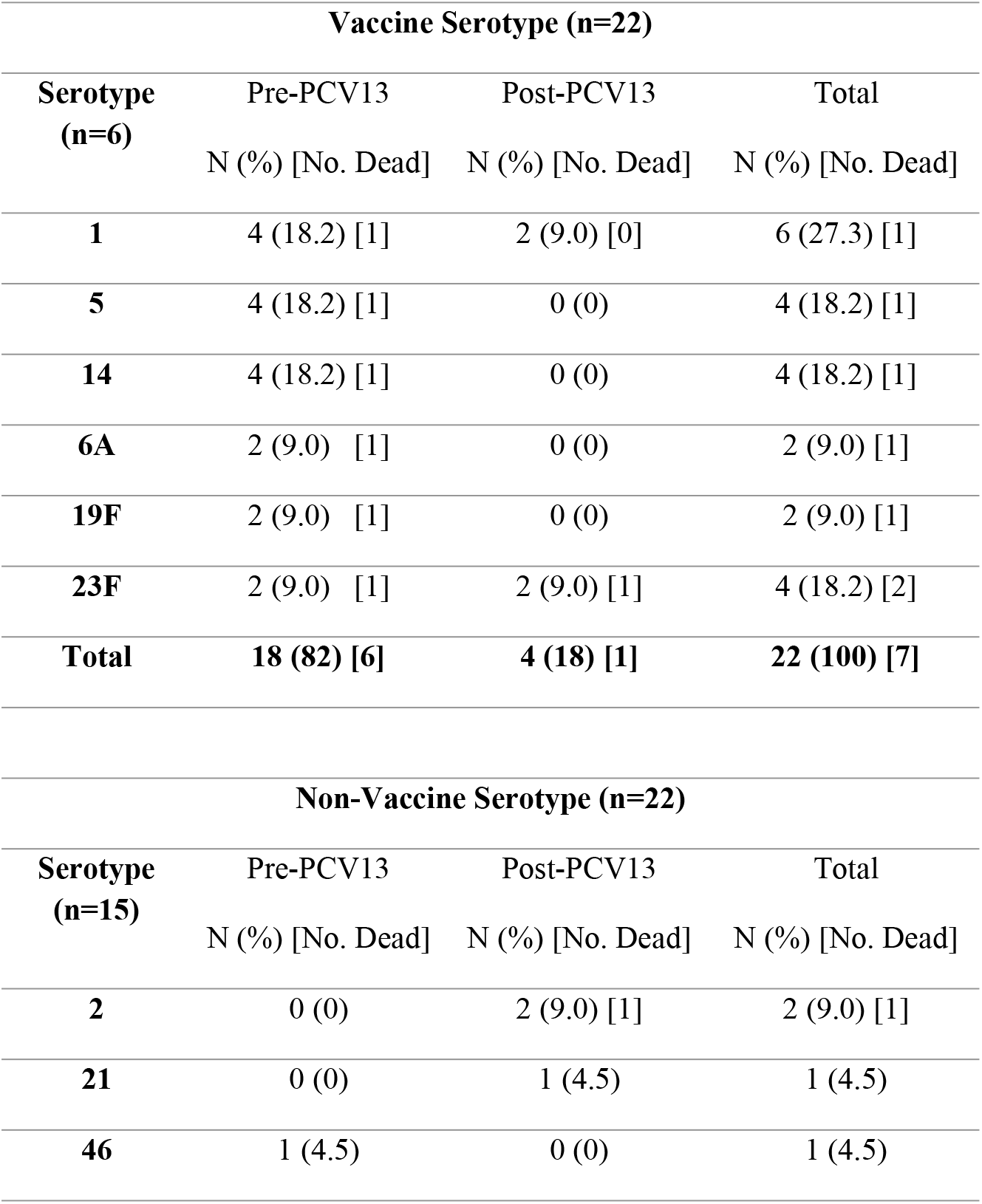
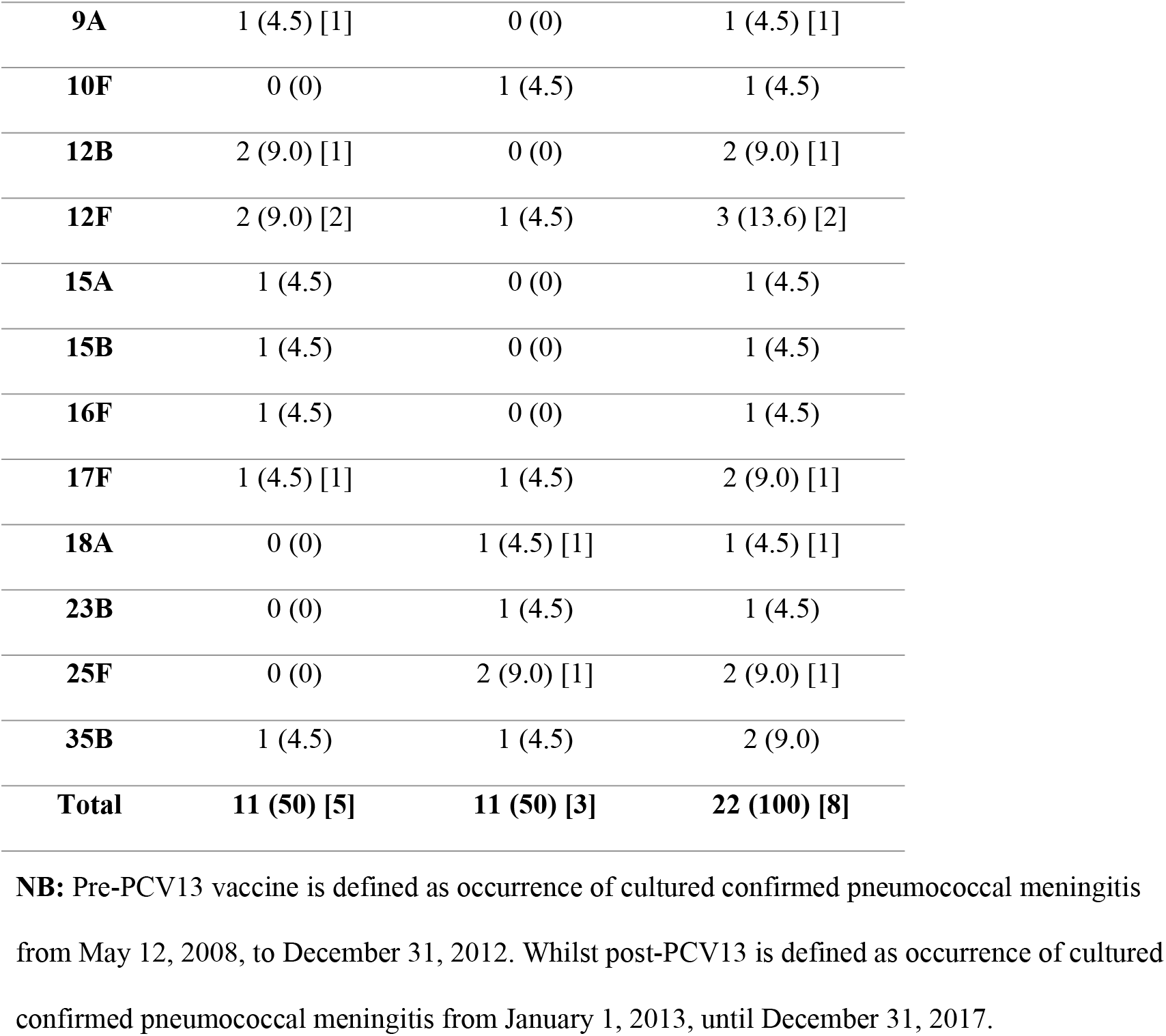
Distribution of Twenty-one Pneumococcal Serotypes Causing Pneumococcal Meningitis (n = 44)

### Bacterial resistance patterns

All pneumococcal isolates were susceptible to penicillin, ampicillin, and cefotaxime with 64% resistant to co-trimoxazole and 14% resistant to chloramphenicol (Table 5). All meningococcal isolates were susceptible to cefotaxime. Resistance among Hib isolates was 8% for cefotaxime, 17% for ampicillin, and 33% for chloramphenicol. *E. coli, K. pneumoniae*, and non-typhoidal Salmonella (NTS) were generally sensitive to cefotaxime, ciprofloxacin and gentamicin but not ampicillin. There were no significant different in the rate of pneumococcal resistance against many antibiotics before introduction of PCV13 compared with post-PCV13. However, the rate of pneumococcal resistance against chloramphenicol and erythromycin was over 100% higher post-PCV13 compared to pre-PCV13.

**Table 5:** Bacterial Antimicrobial Resistance patterns against nine antibiotics

| Bacteria | Number of Isolates | Antimicrobial Resistance, n (%) |  |  |  |  |  |  |  |  |
| --- | --- | --- | --- | --- | --- | --- | --- | --- | --- | --- |
|  |  | AMP | CTX | CHL | CIP | SXT | ERY | PEN | TET | CN |
|  |  | N (%) | N (%) | N (%) | N (%) | N (%) | N (%) | N (%) | N (%) | N (%) |
| *S. pneumoniae* | 44 | 0 (0) | 0 (0) | 6 (14) | 12 (27) | 28 (64) | 4 (9) | 0 (0) | 18 (41) | N/A |
| *N. meningitidis* | 42 | 5 (12) | 0 (0) | 3 (7) | 0 (0) | 27 (64) | 2 (5) | 2 (5) | 4 (10) | N/A |
| Coliform (GNR) | 26 | 7 (27) | 4 (15) | 5 (19) | 3 (12) | 9 (35) | N/A | N/A | 6 (23) | 8 (31) |
| *Staphylococcus aureus* | 16 | *OX<br>5 (31) | N/A | 0 (0) | N/A | 6 (38) | 2 (13) | 12 (75) | 4 (25) | 3 (18) |
| *Hib* | 12 | 2 (17) | 1 (8) | 4 (33) | 0 (0) | 8 (67) | 6 (50) | 3 (25) | 4 (33) | N/A |
| NTS | 12 | 5 (42) | 0 (0) | 1 (8) | 1 (8) | 1 (8) | N/A | N/A | 1 (8) | 1 (8) |
| Non-*Hib* | 6 | 0 (0) | 0 (0) | 3 (50) | 0 (0) | 0 (0) | 1 (17) | 0 (0) | 3 (50) | N/A |
| *K. pneumoniae* | 6 | 1 (17) | 2 (33) | 1 (17) | 0 (0) | 1 (17) | N/A | N/A | 0 (0) | 0 (0) |
| *Escherichia coli* | 5 | 2 (40) | 0 (0) | 2 (40) | 0 (0) | 1 (20) | N/A | N/A | 1 (20) | 2 (40) |

**Key:** AMP Ampicillin, CTX Cefotaxime, CHL Chloramphenicol, CIP Ciprofloxacin, SXT Cotrimoxazole, ERY Erythromycin, PEN Penicillin, TET Tetracycline, CN Gentamycin and OX Oxacillin, GNR Gram-negative rod and N/A Not Applicable
**Note:** Disk diffusion methods were used following standard guidelines (CLSI 2012, M100-S22, Vol. 32 No.3).

## Discussion

This paper provides new information on meningitis incidence and aetiology in The Gambia during a period after the introduction of Hib and pneumococcal conjugate vaccine.

Over the study period, there was steady decrease in the incidence of ABM except in 2012 when there was an outbreak caused by *N. meningitidis* serogroup W [15], as was the case in other parts of the meningitis belt in 2012 and 2015 [16]. The gradual reduction in the incidence of ABM was likely due to the introduction vaccination strategies targeting the three most common bacterial causes of meningitis - *H. influenzae* type-b [17], *N. meningitidis* [18-19] and *S. pneumoniae* [12, 20]. However, the annual incidence of clinically suspected but not confirmed and NBM cases remained relatively stable from 2008 to 2017 with a mean incidence of 141.9, per 100,000 and 26.45, per 100,000 respectively. Few studies in sub-Saharan Africa have reported on cases of NBM [21].

Despite progress in decreasing the mortality caused by ABM in the Gambia, there were still many childhood deaths due to ABM in this rural study, with an overall CFR for ABM of 29% and a CFR of 12.8% for CSM and 8.6% for NBM cases. A similar study in Taiwan showed a CFR 10.9% (6/56) among ABM cases but a CFR of 0% (0/141) among NBM cases [22]. It is unclear whether deaths observed among NBM children in our region were due to infection with other microbial agents, or whether these children had encephalitis as well as meningitis with the former having high mortality. It may also have been due to missed diagnosis cause by prior-antibiotic use before lumber-puncture.

The annual CFR for ABM compared to CSM and NBM was consistently higher throughout the study period. The reason for the particularly high fatality in years 2008, 2010, 2011, 2016 and 2017 is unclear. This observation was also made in earlier studies from Africa and Europe describing the high mortality of bacterial vaccine-preventable diseases[7, 23]. Many previous studies have reported a high CFR for ABM [24] but few have shown a significant level of CFR in CSM and NBM cases. Our study also showed that 75% (126/169) of the ABM related death occurred within seven days of hospitalization whilst the remainder (43/169) occurred within three weeks following hospital discharge.

Both the incidence of ABM and its case fatality rate were highest in the youngest age group in keeping with other studies [15, 25], perhaps due in part to the fact that these children had not been vaccinated; a high incidence and CFR from NBM group was seen also among the youngest children. There are no recent studies from West-Africa that report incidence and CFR of CSM and NBM with which to compare our findings. There was no statistically significant difference of ABM between genders although, the incidence of ABM was proportionately higher among males compared to females, as reported in a similar study in Brazil [25]. A concern is the high incidence in cases of NBM which can cause clinical diagnostic uncertainty, empirical-antibiotic-treatment and undirected patient management. In-turn, this can result in a poor patient outcome, increased risk of child disability[26–28] and a further burden to hospital resources and society [29].

The number of cases of *S. pneumoniae* meningitis due to vaccine serotype decreased substantially 12 months after the introduction of PCV13 vaccination from 82% to 18%. Nevertheless, *S. pneumoniae* was the most common cause of ABM with vaccine serotype and non-vaccine serotype accounting for 50% (22/44) each. Cases of meningitis due to non-vaccine serotypes remained relatively constant in pre-PCV13 and post-PCV13 which strengthens the evidence that the reduction in cases of vaccine serotype meningitis seen after the introduction of PCV was due to the vaccine, as found in earlier studies [12]. Immunisation with Hib conjugate vaccine was introduced into routine EPI programme of The Gambia in 1997 and from 2008 to 2010 there were no bacterial meningitis cases due to Hib in the study area. However, there were sporadic Hib cases from 2011 to 2013 and 50% of all the Hib cases were identified in 2013 and in 2013, a study undertaken in the same region reported an incidence of Hib disease of 88 (95% CI 29-207) per 100,000 in those aged 2-11 month and 22 (95% CI 9-45) per 100,000 in those aged 2-59 months. The reason for this sudden rise in the incidence of Hib was unexplained but cases included a few vaccine failures and many cases in babies too young to have been vaccinated [6]. A recent report indicated that Hib transmission continues at a low rate in The Gambia without a booster vaccination[30]. Our study included several cases of meningitis due to *K. pneumoniae, E. coli* and non-typhoidal Salmonella agreeing with previous studies that have drawn attention to the important role of gram-negative bacilli in ABM [10], [31-32].

We assessed the pattern of bacterial resistance against commonly used and readily available antibiotics in rural Gambia. Our result showed that most bacteria that cause meningitis in the rural Gambia are still highly susceptible to cephalosporin, and cefotaxime was identified as the most effective against gram-negative bacilli. An urban Gambia Invasive Bacterial Disease report showed a similar antibiotic resistance pattern [33].

The findings from our study underscore the urgent need for newer molecular diagnostics[34–36] that better identify viral, bacterial and fungal pathogens, which could guide earlier and more appropriate clinical management of children with meningitis in sub-Saharan Africa. In addition, surveillance against the emergence of antibiotics needs to be sustained.

Although this study is one of the largest to describe ABM, NBM and CSM in west-Africa, the study had several limitations, which included an inability to employ molecular approaches to diagnose the aetiology of meningitis, the absence of viral and fungal diagnoses, a lack of capacity to do detailed follow-up after discharge from hospital to account for either acute or severe neurological sequelae outcome and lack of consideration for assessment of the length of hospitalisation in the analysis.

Overall, findings from this study emphasize the continuing burden of ABM in low income countries like The Gambia despite the introduction of effective conjugate vaccines and the need to support the World Health Organisation recently lunched new meningitis global strategic goals [37] which aims to defeat meningitis by 2030 and to save over 200,000 lives annually.

## Data Availability

Data are available from the Medical Research Council Unit Gambia Institutional Data Access / Ethics Committee (contact via principal investigator) for researchers who meet the criteria for access to confidential data.

## Acknowledgements

We thank the staff of Basse District Hospital, and staff of other health facilities in Upper River Region. We also thank the staff of the Expanded Programme on Immunisation and the Gambia government for their unwavering collaboration with the Medical Research Council Unit at the London School of Hygiene and Tropical Medicine The Gambia. We thank all staff who worked on the Pneumococcal Surveillance Programme and Basse Health Demographic Surveillance System (BHDSS) for their support and the residents living in the area surveyed by the BHDSS team for their participation in the study.

